# Decreased interleukin-8 levels in the peripheral blood of Alzheimer’s patients

**DOI:** 10.1101/2024.12.12.24318919

**Authors:** Francisco Ros, María Dolores Martínez Lozano, Javier Arnau, Jacobo Martínez, Alejandro Vericat, Rafael Sánchez, Javier S. Burgos

## Abstract

**Introduction:** Multiple studies have reported variations in inflammatory levels in blood of Alzheimer’s disease (AD), although further research is needed to identify discriminatory biomarkers for AD diagnosis.

**Methods:** A prospective comparative paired study of a group of AD patients compared to normal controls (NC) was carried out in a multicenter study, where age and sex ratio were homogenized to avoid bias. The cohort comprised 39 patients with a diagnosis of AD (64.1% women; mean age 74.8±0.8 years) and 27 matched NC (66.7% women; 74.8±0.8 years). The diagnosis of AD was performed by neurologists using the Mini-Mental State Examination test (MMSE) and classical markers in cerebrospinal fluid (CSF) and/or amyloid-PET. We investigated the discriminatory potential of 13 serological parameters in blood, consisting of seven interleukins (IL-3, IL-4, IL-5, IL-6, IL-8, IL-12p40, and IL-15) and other 6 serological inflammatory markers (EGF, GM-CSF, IP-10, sCD40L, PDGF-AB/BB, and RANTES). Additionally, six sociodemographic variables were examined. Apolipoprotein E (APOE) genotype was also determined.

**Results:** The results revealed an inverse association between educational level, number of languages spoken, and physical activity, with the risk of AD. An enrichment of the APOE4 isoform was observed in AD (28 out of 39 AD vs 3 out of 24 NC). All circulating inflammation markers were lower in AD patients than in NC (except for IP-10 and sCD40L, while Il-3 and GM-CSF were undetectable in both groups), with statistically significant differences detected in IL-8, IL-12p40, and PDGF-AB/BB (AD/NC ratios of 0.56, 0.59, and 0.81, respectively). IL-8 was decreased specifically in APOE4 carriers (carriers/non-carriers ratio of 0.73), and in amyloid-positive cases (positives/NC ratio of 0.46), whereas IL-12p40 and PDGF-AB/BB showed positives/NC ratios of 0.56 and 0.83, respectively.

**Discussion:** Serum levels of IL-8, IL-12p40, and PDGF-AB/BB were significantly reduced in AD patients. Additionally, IL-8 decrease was associated with APOE4-carriers and amyloid-positive cases.

## 1. INTRODUCTION

Alzheimer’s disease (AD) is the most common progressive and neurodegenerative disease with a dementia component in older adults and is characterized by a mental decline with loss of cognitive skills and memory functions^1^. Pathophysiological senile plaques formed by Aβ peptides, and neurofibrillary tangles of aggregates of hyperphosphorylated tau have long been suggested as the main causes of AD. While these factors are well established, it is evident that AD is a complex disease involving multiple mechanisms, including inflammation^2,3^. Substantial evidence supports the role of inflammation in AD, although the precise relationship between systemic and central inflammation remains unclear.

Given the preclinical nature of AD, early interventions with disease-modifying drugs could potentially delay disease onset. The recent Food and Drug Administration (FDA) approvals of lecanemab in 2023 ^4^ and donanemab in 2024 ^5^ underscore the need for discriminatory and predictive biomarkers to facilitate clinical follow-up and evaluation of these and other potential treatments^4,5^. Prioritization of readily quantifiable, minimally invasive biomarkers is essential for the evaluation of disease risk. Circulating markers would enable widespread application and repeated evaluations, addressing the limitations of current diagnostic methods, which often involve complex and invasive procedures. Traditional AD diagnosis relies on neuropsychological assessments and canonical markers in cerebrospinal fluid (CSF) or PET scans.

In the last two decades, significant efforts have been made to discover new non-invasive markers with discriminatory capacity for AD. Neuroinflammation-related markers have gained special prominence, particularly following the seminal work of Ray et al. in 2007^6^. This study identified 18 signaling proteins in blood plasma that could discriminate between Alzheimer’s and control subjects, including cytokines, chemokines, and growth factors.

Cytokines, including interleukins and chemokines, mediate neuroinflammation responses^7–9^, wherein microglia and astrocytes play a crucial role in modulating pro- and anti-inflammatory molecules^10^. In AD, there is evidence linking neuropathological hallmarks to cytokine-mediated glia activity^11–13^. Several studies suggest that cytokine imbalance precedes the clinical onset of AD^14^, indicating the potential impact of peripheral cytokines on disease progression^15^.

In this study, we assessed a panel of inflammatory proteins in the blood of sex- and age-matched cognitively unimpaired older adults (normal controls [NC]) and individuals diagnosed with AD using MMSE, amyloid, and tau markers in CSF and/or β-amyloid PET. Our objective was to identify novel circulating markers that could differentiate between cases and controls as well as potential targets for disease-modifying therapies.

Following the identification of differentially expressed markers, we investigated the influence of apolipoprotein E genotype and amyloid positivity, finding a direct association between IL-8 and both factors.

## 2. METHODS

### 2.1. Study participants

Participants aged 65 and older were recruited in 2023 from three Spanish clinical centers (Hospital de la Magdalena, Hospital Arnau de Vilanova, and FISABIO). Inclusion and exclusion criteria, disseminated to participating hospitals, guided recruitment. Independent, specialized neurologists at each site assessed all diagnoses based on clinical history, cognitive evaluation, cerebrospinal fluid biomarkers, and/or amyloid PET imaging. Exclusion criteria included psychiatric disorders and cognitive impairments attributable to other systemic or central nervous system conditions.

Control subjects were recruited concurrently from the same geographical area (Comunitat Valenciana, Spain), directly from participating hospitals or the Biobank for Biomedical and Public Health Research of the Comunitat Valenciana (belonging to the FISABIO Foundation). These individuals reported no subjective symptoms of cognitive dysfunction and were recruited using the same exclusion criteria and age requirement.

In addition to age and sex, the following parameters were collected: educational level (no studies, elementary school, middle school, or bachelor’s degree), tobacco use (smokers or non-smokers), alcohol use (≥1 time per week), number of spoken languages, body mass index (BMI), and physical activity (sedentary or regular exercise).

The final case-control study included 39 patients with AD (25 women and 14 men) and 27 age- and sex-matched normal subjects (18 women and 9 men) (Table 1).

**TABLE 1.** Sociodemographic factors of the study cohort.

|  |  | NC (n=27) | AD (n=39) |
| --- | --- | --- | --- |
| Age (years) | Mean | 74.8 ± 0.8 | 74.8 ± 0.8 |
|  | Range | 66.0 - 83.0 | 63.0 - 84.0 |
| Sex (% women) |  | 66.7 | 64.1 |
| BMI |  | 27.6 ± 0.8 | 25.0 ± 0.5 |
| Education level | No studies | 0.0 % | 23.1 % |
|  | Elementary School | 44.4 % | 51.3 % |
|  | Middle School | 33.3 % | 15.4 % |
|  | Bachelor's degree | 22.3 % | 10.2 % |
| Languages spoken | 1 | 34.6 % | 61.5 % |
|  | 2 | 38.5 % | 38.5 % |
|  | 3 | 26.9 % | 0.0 % |
| Tobbaco use |  | 0.0 % | 2.6 % |
| Alcohol use |  | 44.4 % | 5.3 % |
| Physical activity |  | 92.0 % | 50.0 % |

### 2.2. Ethical considerations

The study protocol was approved by the Ethics Committees of Universitat Jaume I (ethical approval number CEISH/44/2022), and the FISABIO Foundation (ethical approval number CEI-SP 20230127/03). Neurologists provided written information and explained the study protocol to the controls, patients and/or caregivers.

### 2.3. Consent statement

Informed consent was obtained from all participants or their guardians in cases where the patient’s capacity to consent was compromised. This study was conducted in accordance with the Declaration of Helsinki.

### 2.4. Clinical examination

Prior to the test day, cognitive assessments were conducted by independent Neurology teams at participating hospitals. Body weight was measured to the nearest 0.1 kg, body height was measured barefoot to the nearest 0.01 m, and body mass index (BMI) was calculated. CSF sampling and amyloid PET analysis were performed as part of routine clinical practice and utilized to determine the final diagnosis.

### 2.5. Blood collection and biochemical procedures

Blood samples were collected via venipuncture from fasted patients on the morning of the test day. To minimize inter-subject variability due to physiological diurnal fluctuations in immune function, patients were seated in a relaxed position, and extractions were performed between 8.00 and 10.00 am. After a 15-30 min coagulation period at room temperature, the samples were processed to obtain serum by centrifugation in pre-treated tubes. Serum aliquots were stored at-80°C for biochemical analyses and were not subjected to thawing and refreezing. Previously, an aliquot of blood was stored in EDTA collection tubes for APOE genotyping.

Biochemical analyses were conducted by a laboratory analyst who was blinded to clinical diagnoses and other patient information. Biomarker quantification was performed at the Hospital Doctor Peset in Valencia, Spain. Serum samples were analyzed using a commercially available Cytokine Human 48-plex panel (MILLIPLEX® Human Cytokine/Chemokine/Growth Factor Panel A-Immunology Multiplex Assay) for the following analytes: EGF, GM-CSF, IL-3, IL-4, IL-5, IL-6, IL-8, IL-12p40, IL-15, IP-10, PDGF-BB, sCD40L (reference: HCYTA-60K-12 Human Cyto Panel A), and RANTES (reference: HCYTA-60K-01 Human Cyto Panel A). Samples were analyzed in duplicate, according to the manufacturer’s recommendations, using the antibody bead mix with biotinylated detection antibody, followed by streptavidin-phycoerythrin. The assay was performed on a Luminex platform (Milliplex), and data were collected for 100 beads per cytokine from each well. Cytokine concentrations were calculated using Milliplex Analyst software, employing a five-parameter curve-fitting algorithm for standard curve fittings. All analyses were conducted in a single batch to minimize inter-assay variability.

### 2.6. APOE genotyping

APOE genotyping was performed on genomic DNA extracted from peripheral blood lymphocytes, as previously described^16^. Forward (5’-cggaactggaggaacaactg-3’) and reverse (5’-ggatggcgctgaggaggccgcgctc-3’) primer sets were designed to amplify *APOE* exon 4 in the genomic DNA samples. This resulted in a 306 bp amplicon that containing c.388 and c.526 positions.

Polymerase chain reaction (PCR) was performed using Firepol DNA polymerase (SolisBioDyne), according to the manufacturer’s protocol. The reaction mixture included primers (10 μM) and 1 μL of 100% dimethyl sulfoxide (DMSO) to facilitate amplification of the GC-rich target region. PCR was performed using a ProFlex PCR system (Applied Biosystems). The thermal cycling program comprised an initial denaturation at 95°C for 2 minutes, followed by 35 cycles of denaturation at 95°C for 30 seconds, annealing at 57°C for 30 seconds, and extension at 72°C for 30 seconds. A final extension step was performed at 72°C for 7 minutes.

Prior to Sanger sequencing, unincorporated primers and nucleotides were removed using an Illustra ExoProStar 1-STEP Kit (Cytiva). Bidirectional Sanger sequencing was performed to determine the presence of the polymorphisms c.388T>C and c.526C>T in the patient samples. Forward and reverse Sanger sequencing were performed to determine the presence of c. 388T > C and c. 526C > T polymorphisms in patients.

### 2.7. Statistical analyses

The study did not use a priori power analysis to determine the sample size. Nonetheless, the sample size was considered sufficient to fulfill the research objectives. Data were extracted from individual files and imported into Jeffreys’ Amazing Statistical Package (JASP; https://jasp-stats.org/) for statistical analysis and visualization. Outliers were identified using a robust regression and outlier method (ROUT, Q=0.1%). Prior to each statistical analysis, the normality of the dependent variable’s distribution was assessed using the Kolmogorov-Smirnov (KS) test, and homogeneity of variances was evaluated using Levene’s test.

The analysis began with descriptive statistics to characterize both groups. A significance level of α < 0.05 was adopted. Independent-samples t-test were employed for between-group comparisons.

All quantitative variables were depicted using raincloud plots, which combined a cloud of points, box plot, and one-sided violin plot. The cloud of jittered points represents individual observations. The box plot illustrates the median (thick line), interquartile range (box), and the minimum and maximum values (error bars). The one-sided violin (density) plot displays the data distribution.

## 3. RESULTS

### 3.1. Sociodemographic data

A comprehensive analysis of sociodemographic factors was conducted to characterize the study cohort (Table 1). The final case-control study included 39 AD patients and 27 age-matched NC. The mean age was 74.8±0.8 years for both groups (ranges of 66.0-84.0 and 63.0-83.0 years, respectively). Women were overrepresented in both patients (64.1%) and controls (66.7%), aligning with the previously described sex-biased vulnerability in AD^17–19^, demonstrating perfect pairing between both groups. BMI was also similar between the groups but significantly lower in patients (p = 0.008).

Regarding cognitive reserve factors, educational level was inversely associated with AD prevalence. Indeed, assigning values from 0 to 3 (0: no education, 1: elementary school, 2: middle school, and 3: bachelor’s degree), the score was significantly higher (p=0.007) in the NC group than in the AD patients (1.78 versus 1.13; a decrease of 36.5%). Additionally, multilingualism exhibited a protective effect against AD, with 61.5% of the AD patients speaking only one language and 65.4% of the NC speaking two or more languages, indicating a significant difference between groups (p= 0.02).

Concerning toxic habits, only one subject in the entire cohort was a smoker (an AD patient). In contrast, the NC group exhibited significantly higher alcohol consumption habits than patients (44.4% vs. 5.3%; p= 0.03), suggesting a more active social life among controls. This discrepancy may be attributed to the increased dependence of AD patients. Moreover, control subjects had substantially higher physical activity levels than patients (92.0% vs. 50.0%; p< 0.001), likely due to similar reasons.

### 3.2. APOE genotyping

Statistical analysis of the APOE genotypic distribution revealed significant differences between NC and AD patients (Figure 1). Patients were predominantly carriers of at least one copy of the APOE4 allele (28 out of 39 cases; 71.8%), whereas controls presented a much lower frequency of this isoform (3 out of 27 controls; 11.1%), confirming previous findings of APOE4 enrichment in AD patients^19^. The predominant genotypes for controls were APOE3/3 (23 out of 27; 85.2%) and APOE3/4 for patients (24 out of 39; 61.5%).

**FIGURE 1.**
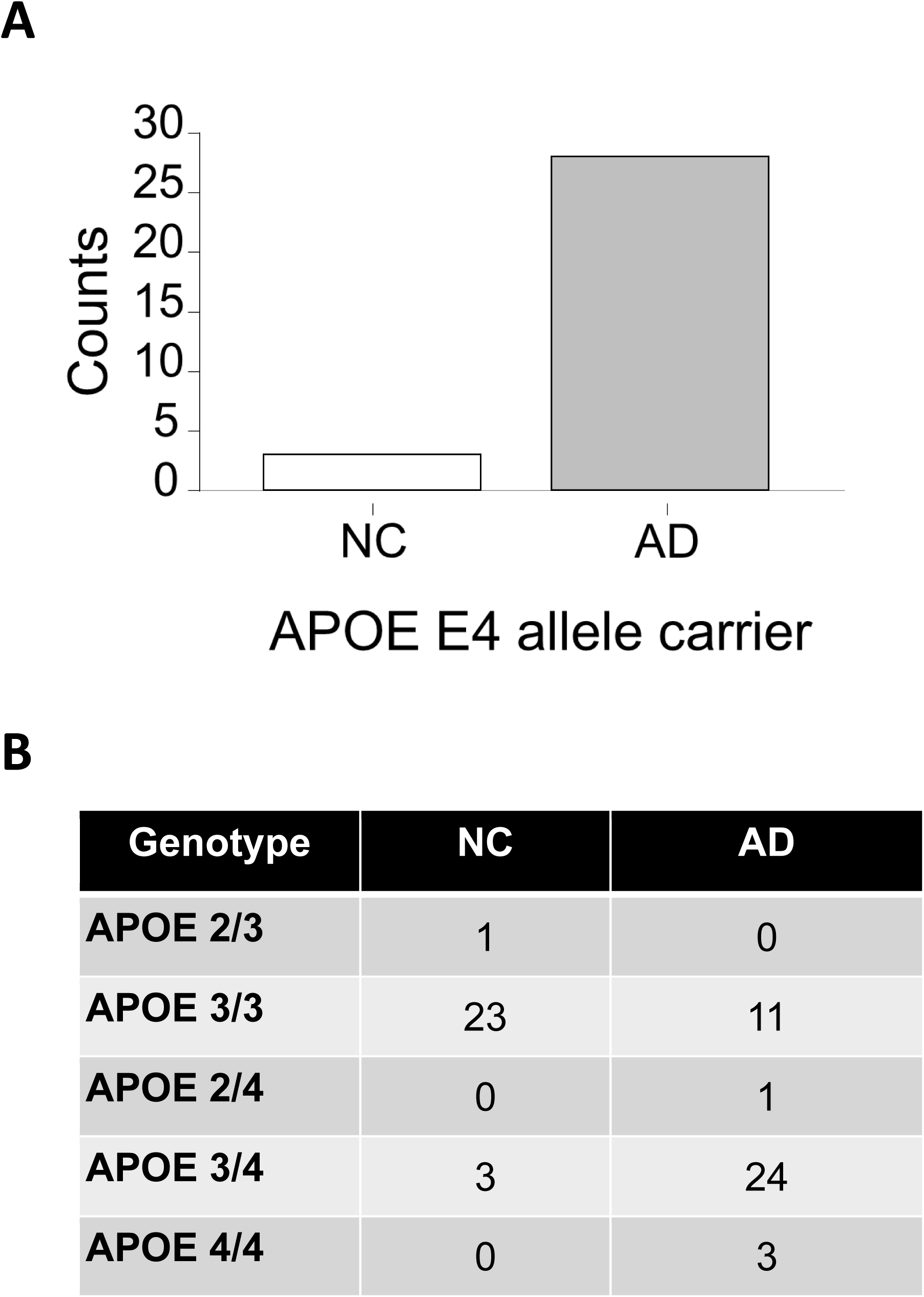
APOE genotypic distribution within the study cohort. A bar graph illustrates the number of control and AD individuals carrying at least one E4 allele (A), while a table presents the allelic frequency distribution of APOE variants across both groups.

### 3.3. Serum concentrations of markers

Protein concentrations were evaluated in the serum of NC and AD patients (Figure 2). IL-4, IL-5, IL-6, IL-8, IL-12p40, IL-15, IP-10, EGF, PDGF-AB/BB, sCD40L, and RANTES were detectable within the assay’s dynamic range in serum, while IL-3 and GM-CSF were below the limit of detection; therefore, they were not reported. In the patient group, serum concentrations of all inflammatory markers decreased (except for IP-10 and sCD40L), although only the reductions in IL-8 (p<0.001), IL-12p40 (p<0.001), and PDGF-AB/BB (p= 0.004) were statistically significant.

**FIGURE 2.**
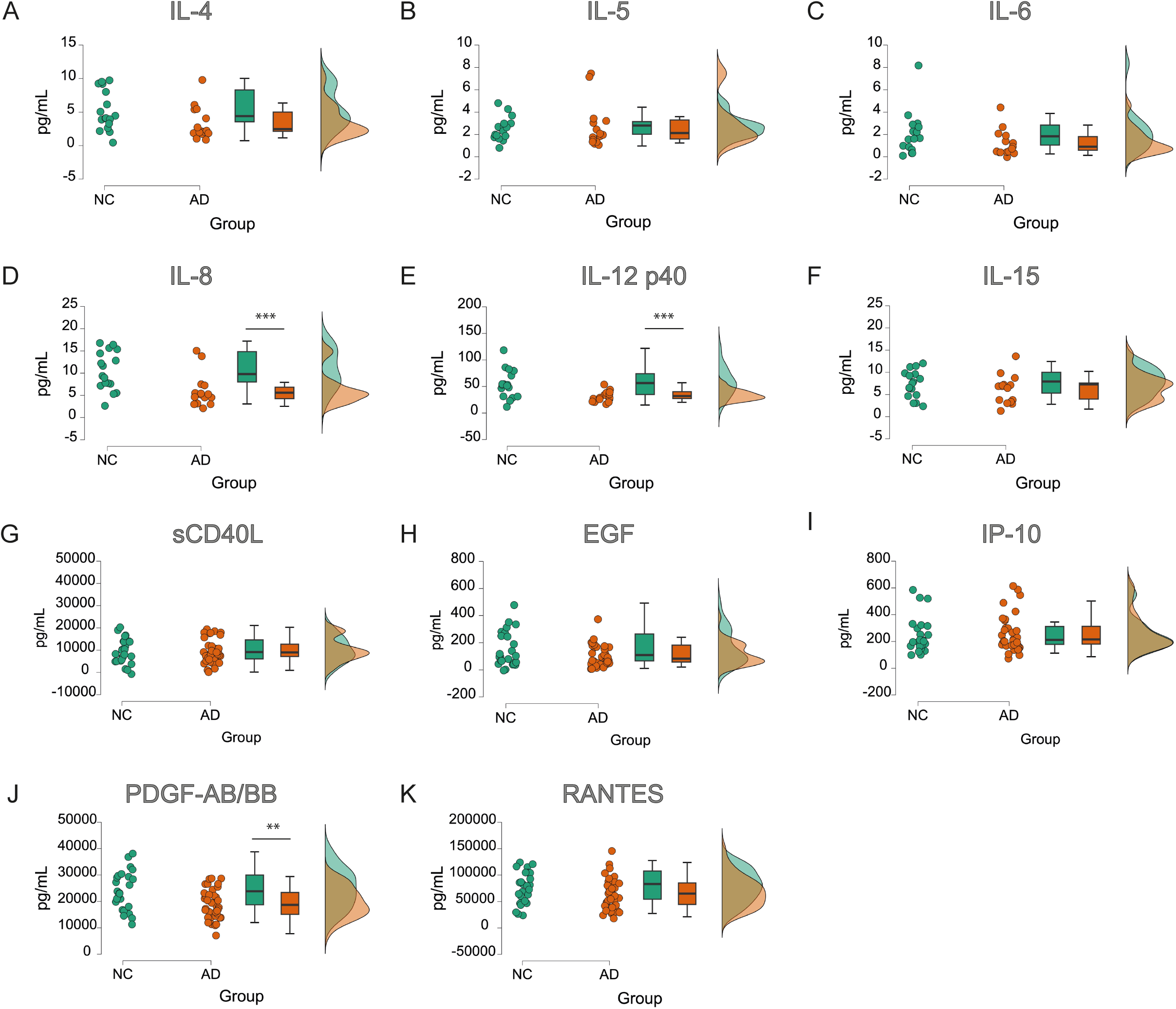
Protein concentrations of the different evaluated markers in NC and AD subjects. Violin plots were used to visualize the data distribution and compare concentrations between the two groups. Results revealed statistically significant differences in the levels of IL-8 (D, ***), IL-12p40 (E, ***), and PDGF-AB/BB (J, **) between groups. A significance level of 0.05 was set, and independent sample t-tests were performed. **p≤0.01; ***p≤0.001).

In an additional analysis, the subjects were divided into carriers and non-carriers of APOE4, regardless of group affiliation, and inflammatory markers were compared again (Figure 3). All tested proteins exhibited lower levels in the APOE4 carrier group (n=31) than in the non-carriers (n=35) (except for sCD40L), although only IL-8 showed significant differences (p=0.01) in this analysis.

**FIGURE 3.**
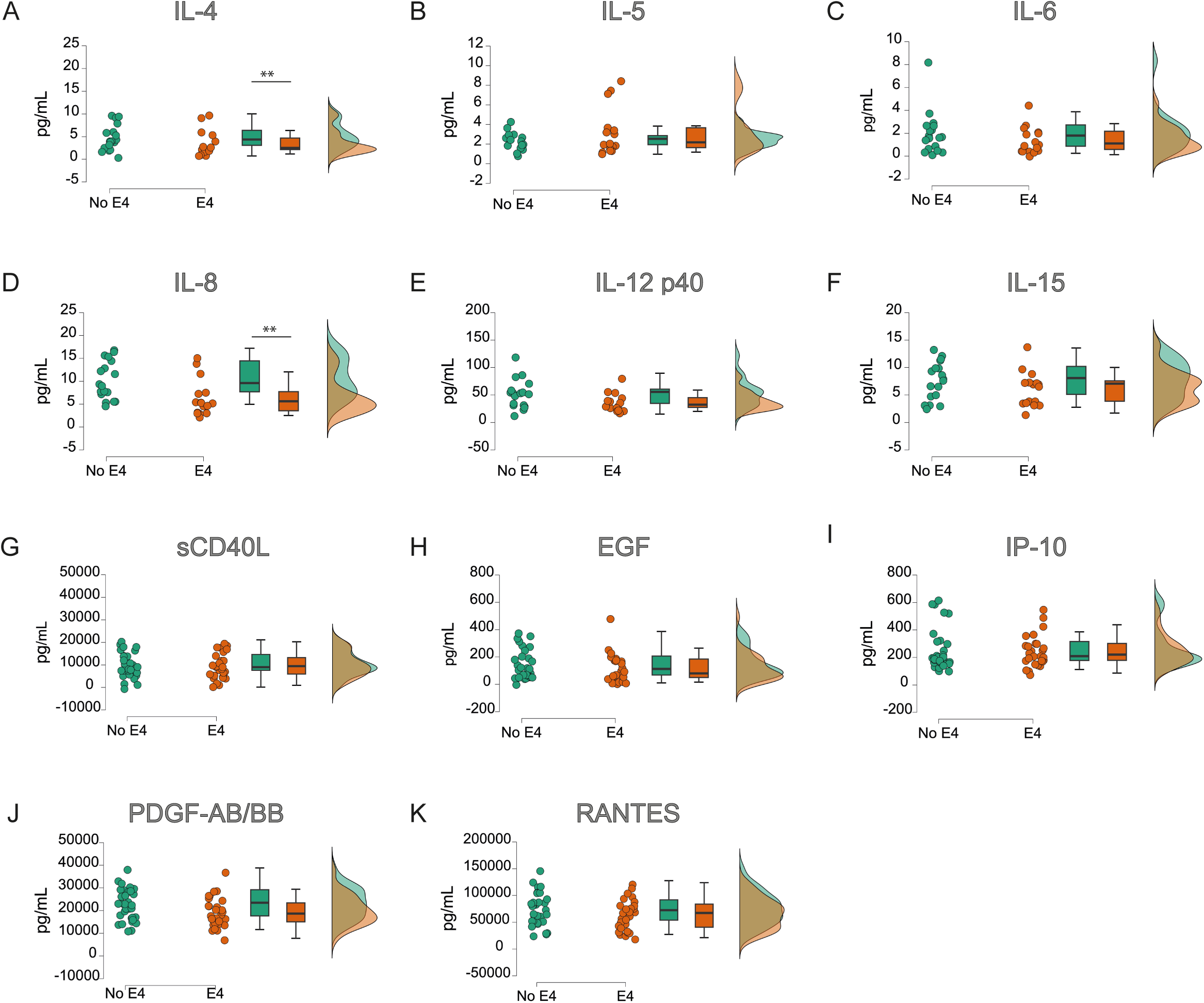
Protein concentrations of the different evaluated markers in APOE4 carriers and non-carriers. Violin plots were used to visualize the data distribution and compare concentrations between the two groups. Results revealed statistically significant differences in the levels of IL-4 (A, **) and IL-8 (D, **) between groups. A significance level of 0.05 was set, and independent sample t-tests were performed. **p≤0.01.

Finally, the three proteins with significant differences between AD patients and NC (IL-8, IL12p40, and PDGF-AB-BB) were compared for amyloid positivity (determined by CSF and/or PET) (Figure 4). Significant differences between amyloid-positive (n=23) and amyloid-negative (n=27) samples were detected in all cases (IL8: p<0.001; IL12: p= 0.001; PDGF: p= 0.04).

**FIGURE 4.**
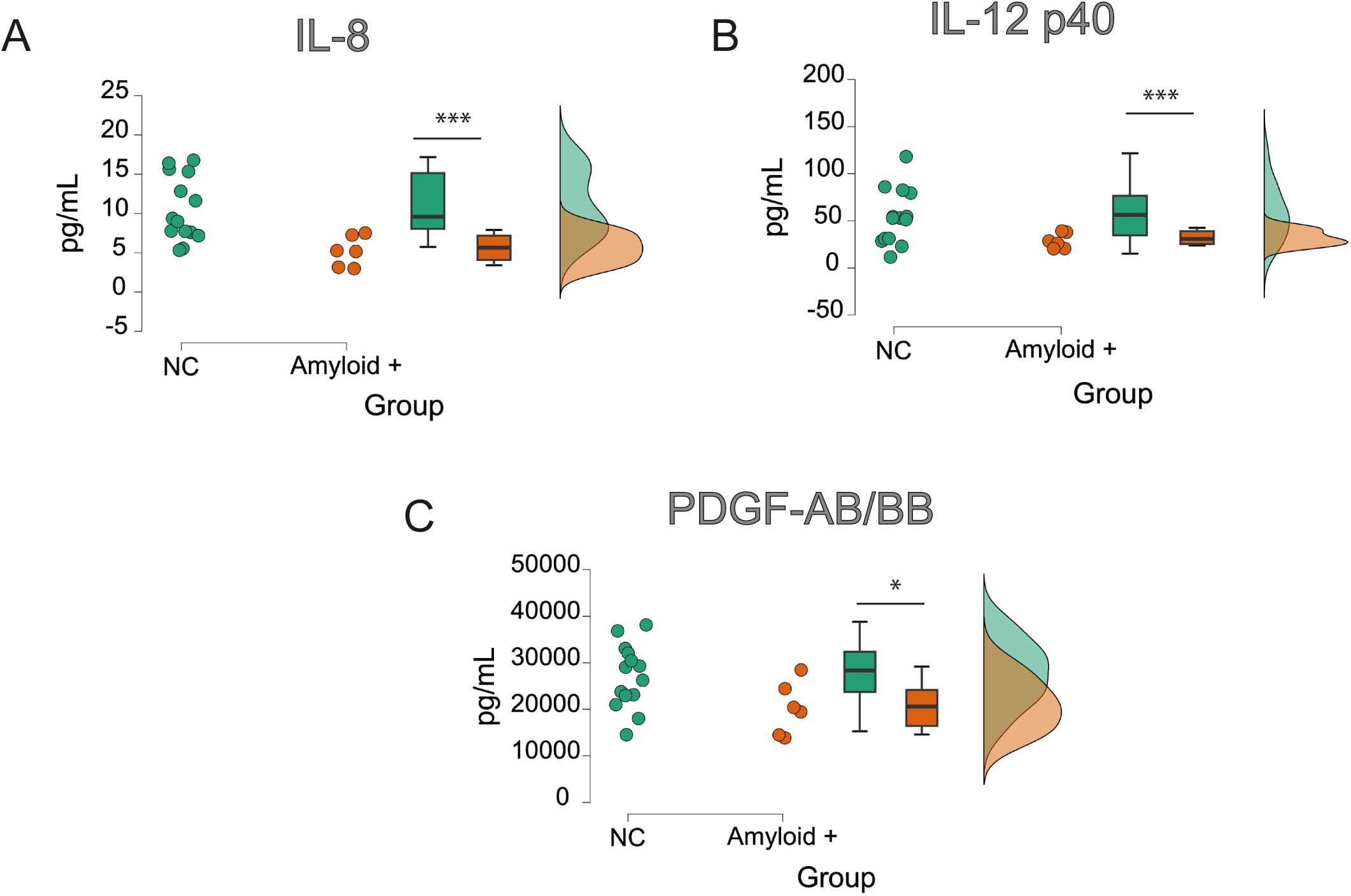
Protein concentrations of IL-8, IL-12p40 and PDGF-AB/BB in NC compared to amyloid-positive cases. Violin plots were used to visualize the data distribution and compare concentrations between the two groups. Results revealed statistically significant differences in the levels of IL-8 (A, ***), IL-12p40 (B, ***), and PDGF-AB/BB (C, *) between groups. A significance level of 0.05 was set, and independent sample t-tests were performed. *p≤0.05; ***p≤0.001.

After analysis of IL-8, IL12p40, and PDGF-AB/BB levels, we studied the different ratios related to the analyzed conditions of the cohort, observing decreasing AD/NC ratios of 0.56, 0.59 and 0.81, respectively (Figure 5A). When the cohort was divided between APOE4 carriers and non-carriers only IL-8 showed significant differences, with a carrier/non-carrier ratio of 0.73 (Figure 5B). Finally, the comparison between amyloid-positive subjects versus NC subjects showed ratios of 0.46 for IL-8, 0.56 for IL12p40, and 0.83 for PDGF-AB/BB (Figure 5C).

**FIGURE 5.**
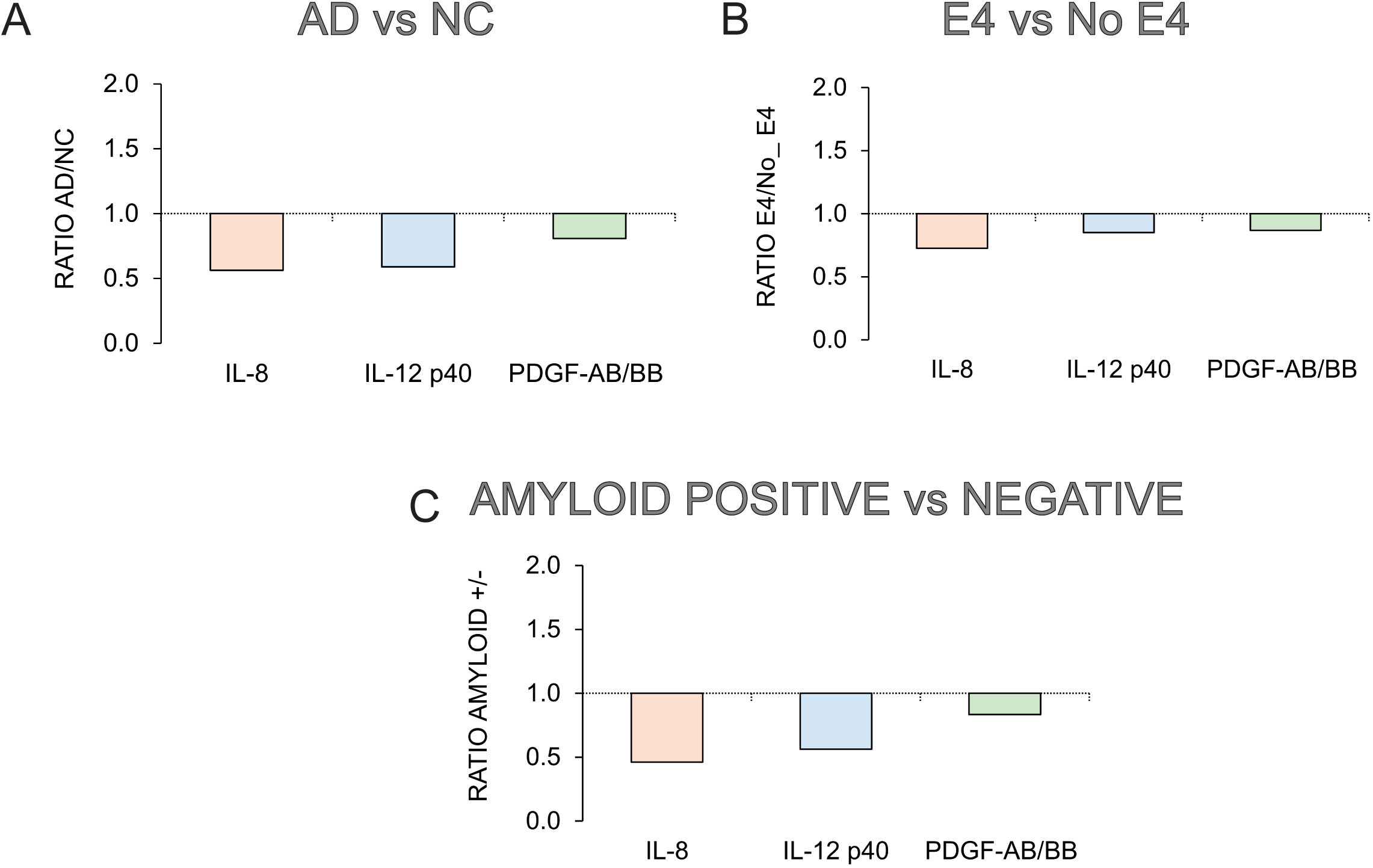
Protein concentration ratios for IL-8, IL-12p40, and PDGF-AB/BB. Bar graphs illustrate these ratios when comparing AD to NC groups (A), APOE4 carriers to non-carriers (B), and amyloid-positive to NC (C).

## 4. DISCUSSION

While amyloid-beta (Aβ) plaques and tau neurofibrillary tangles remain quintessential hallmarks of AD, over the past two decades the inflammatory component, both central and peripheral, has gained prominence among the events related to AD pathogenesis. A substantial body of work has underscored the significance of peripheral immune alterations, particularly proinflammatory signaling, in AD development. Indeed, peripheral inflammation has been identified as a potential risk factor for AD and vascular dementia^20,21^. Cross-sectional analyses and cohort studies have consistently shown that cognitively normal individuals with elevated inflammatory markers in the blood are at increased risk of future mild cognitive impairment, AD dementia, and all-cause dementia^22–26^. These findings highlight the need for further research to elucidate how disease stage, underlying pathology, and specific inflammatory molecules may influence the relationship between inflammatory proteins and AD progression.

In the present case-control study, we analyzed a series of inflammatory markers at the systemic level and determined their variations in patients and controls, segregating them by APOE genotype and Aβ positivity. In this multicenter study, patients and NC were strictly matched in terms of age and proportion of women, with the aim of reducing heterogeneity between the groups and drawing accurate conclusions about the circulating biomarkers under study. Moreover, patients with AD were enriched in the APOE4 allele compared to NC, confirming the classical association of this genotype with the disease^19^. Education level was inversely related to AD, and multilingualism was also shown to have a protective effect against AD.

A series of proteins related to inflammatory processes were analyzed with respect to the determination of markers in blood. This strategy was chosen with the aim of identifying new non-canonical markers to discriminate between patients with AD and NC and to deepen the understanding of their implications in the course of the disease. Blood-based biomarkers may be more attractive than classic biomarkers, such as CSF tau and Aβ levels or brain amyloid imaging, because these techniques are invasive, time-consuming, and/or expensive. A convenient blood test to detect AD based on circulating markers would be a valuable tool to add to the diagnostic arsenal for clinicians, at least in the first line of evaluation. Currently, the diagnosis of AD requires a combination of neuropsychological testing, imaging techniques, CSF analysis after lumbar puncture, and the exclusion of other neurological disorders. Moreover, the identification of new blood markers is of particular interest in the current context, especially following the recent FDA approvals of lecanemab and donanemab^4,5^, as the identification of minimally invasive markers will be most useful for screening in large-scale clinical trials and for clinical use. Blood-based biomarkers are not yet routinely used in clinical practice but may be more useful because they are easily obtained with a lower risk of complications in older adults with comorbidities.

To elucidate the role of inflammation in AD, we conducted a comprehensive analysis of up to 13 proteins associated with the inflammatory response in the serum of patients and normal controls using a Milliplex™ panel of cytokines. While two of them were undetectable in either group, two others showed no difference between groups, and the remaining nine exhibited lower levels in patients than in controls. Notably, three of these proteins (IL-8, IL12p40, and PDGF-AB/BB) demonstrated statistically significant reductions in patients, with a decreasing order of modulation.

Interleukin-8 (IL-8), a chemokine produced by macrophages in response to proinflammatory mediators such as amyloid, may play a critical role in recruiting activated microglia to sites of AD brain damage^27^. Indeed, localization of the IL-8 receptor (CXCR2) in dystrophic neurites suggests a potential mechanism for IL-8-mediated glial interactions with neurons, thereby contributing to neuronal damage^28^. While previous studies have reported significantly elevated IL-8 levels in the CSF of AD patients compared to controls^29^, plasma levels in late-onset AD have shown inconsistent findings, with some studies reporting no difference between patients and controls in European populations^30^. A meta-analysis, limited by the small number of studies examining IL-8, did not find conclusive evidence of its involvement in AD^31^. However, other studies have demonstrated increased IL levels in AD^32^. Consistent with our results, Kim et al. observed significant lower IL-8 concentrations in patients with MCI or AD compared to controls^33^. Furthermore, Shen et al. reported diminished peripheral IL-8 levels in AD patients compared to those with mild cognitive impairment^34^.

Interleukin-12 (IL-12), a pleiotropic and heterodimeric cytokine implicated in bacterial infections and autoimmune diseases^35,36^, is composed of a 35 kDa light chain (the p35 subunit encoded by *IL12A*) and a 40 kDa heavy chain (the p40 subunit encoded by *IL12B*)^36^. Single nucleotide polymorphisms (SNPs) in the genes encoding *IL-12A* (rs2243115 and rs568408) and *IL-12B* (rs3212227) have been linked to the risk of AD in a Chinese population, suggesting their potential role as risk factors for late-onset AD susceptibility^37^. In our study, we observed lower serum levels of IL-12p40 in AD patients. In this sense, previous research has demonstrated a correlation between IL-12 levels and AD severity, with elevated levels in mild to moderate AD and decreased levels in severe AD^38^. In this regard, Yang and co-workers in a study analyzing nine cytokines found that higher levels of IL-12p70 were associated with slower cognitive decline in AD^39^. Additionally, Swardfager et al. reported increased IL-12 levels in the blood of patients with mild and moderate AD^40^, whereas severe AD was associated with suppressed plasma IL-12 levels^38^. These findings align with our results and suggest a potential role of IL-12 in AD progression.

Human platelet-derived growth factor (PDGF) is a potent mitogen for several cellular types, including vascular smooth muscle cells and brain glial cells, and is a primary mitogen in platelet-derived human serum^41^. The PDGF family consists of four members (A, B, C, and D), each formed by disulfide-linked homo- or heterodimers of two distinct but related chains. PDGF binds to specific cell membrane receptors, activating a tyrosine-specific kinase that catalyzes the autophosphorylation and subsequent phosphorylation of cellular proteins^42^. In addition to its mitogenic functions, PDGF regulates diverse cellular metabolic processes, including protein, lipid, and prostaglandin synthesis^43^. While PDGF C and D were identified in the early 2000s, the predominant forms of PDGF are homodimeric or heterodimeric combinations of A and B chains, resulting in PDGF AA, BB, and AB. Notably, AB and BB dimers, but not AA dimers, have demonstrated significant angiogenic properties^44^. Additionally, the PDGF B-chain has been observed to be upregulated in neurons and macrophages of rat brains following cerebral ischemia^45^. Recently, Mizutani et al. demonstrated that Aβ markedly decreased the secretion of PDGF-AB into the plasma by thrombin-activated human platelets^46^. Regarding AD, Ray et al. identified PDGF-BB as a pivotal protein in a plasma signature differentiating AD from controls^6^. Their study revealed lower PDGF-BB expression in AD subjects than in controls, corroborating our findings. Also, Aβ has been shown to inhibit the neuroprotective PDGF-BB system, potentially contributing to the pathogenesis of AD^47^. Additionally, loss of the PDGF-BB receptor in AD has been linked to fibrinogen leakage, reduced oxygenation, and accumulation of fibrillar Aβ^48^. Collectively, these studies emphasize the robust association between PDGF-AB/BB and AD, underscoring its potential significance for patient stratification.

In summary, numerous peripheral cytokines have been identified in AD studies, exhibiting variations at different disease stages. Peripheral blood mononuclear cells presented elevated cytokine levels in patients with MCI compared to those in the AD or controls group^49^, suggesting that inflammatory marker modulation is influenced by disease progression. Notably, Koyama et al. observed a correlation between increased levels of circulating inflammatory markers and a higher risk of dementia later in life, suggesting that peripheral inflammation may precede clinical symptoms^26^. In this regard, studies primarily conducted in patients with early-stage AD and MCI generally report increased plasma and CSF levels of cytokines and chemokines, while decreased plasma cytokine levels have been implicated in altered immune response in moderate-stage AD patients^50^.

Among the strengths of this study, is its representative cohort, which aligns with the sociodemographic characteristics typically observed in AD. Additionally, the study groups were perfectly matched for sex and age, and the APOE genotyping results were consistent with the prevalence of APOE4 carriers among AD patients. Another strength lies in the marker determination system; the multiple bead-based detection assay offers clear advantages over more sophisticated methods, as it can easily and simultaneously detect numerous analytes using minimum sample volumes.

However, this study also has certain methodological caveats and limitations. The most significant limitation was the small sample size despite the statistical robustness of the results. Furthermore, the cross-sectional design of the study precluded longitudinal analysis of the discriminatory markers.

In any case, blood-based biomarkers hold promise for future clinical practice in AD due to their affordability, accessibility, and minimally invasive nature. Therefore, they have the potential to become valuable first-line diagnostic tools. Moreover, the combination of blood biomarkers of AD pathology and neurodegeneration (such as pTau-181 and NF-L) with other non-canonical markers can enhance patient stratification, aiding a better prediction of cognitive impairment and disease stage classification.

## Data Availability

All data produced in the present study are available upon reasonable request to the authors

## ACKNOWLEDGEMENTS

We would like to express our sincere gratitude to the patients and their caregivers, as well as to the control group participants, for their invaluable contributions to this study. We also acknowledge the institutional support of the Hospital de La Magdalena, the Hospital Arnau de Vilanova, the Hospital General Universitario de Castellón, and the FISABIO-Public Health Unit of the IBSP-CV Biobank (PT23/00034), which is part of the ISCIII Biomodels and Biobanks Platform and the Valencian Biobanking Network. Furthermore, we are deeply grateful for the generous donation of Paula Batet and Pau Torres, which made this research possible.

